# Interpreting the result of a diagnostic test: An experimental study in a social network

**DOI:** 10.1101/2020.07.12.20151910

**Authors:** Ahmad Sofi-Mahmudi

## Abstract

**Background:** Statistical literacy is an important element in informing patients and planning treatments for both clinicians and policymakers. This study aim to investigate and compare the interpretation of a diagnostic test result in medical and non-medical professionals.

**Methods:** A diagnostic test question about positive predictive value was posted on the “Stories” section of five personal Instagram accounts. The collected information included the exact calculated number, educational status (undergraduate or graduated) and field of study (dentistry, medicine, other medical, non-medical). The results were analysed using R version 3.4.4 on Microsoft Windows 10 with the Pearson χ2 test.

**Results:** Of 121 participants, 63.6% did not provide a correct answer. The highest and the lowest correct answer ratios were seen in non-medical and dentistry groups, with 52.6% and 22.2%, respectively (P = 0.09, χ2 = 6.464, df = 3). Undergraduates showed a more favourable performance than graduates with 41.8% correct answers compared to 20.0% (P = 0.03, χ2 = 4.616, df = 1).

**Conclusion:** The statistical interpretation of the medical professionals is lower than the non-medical professionals and graduates showed less favourable results compared to undergraduates. There is a need for more beneficial and continuative statistical education for medical professionals.

## Introduction

Statistical literacy is a concern for patients, practitioners, and policymakers. Inappropriate interpretation of health outcome statistics can cause misunderstandings that may lead to preventable unfavourable health outcomes (1). For example, by under- or over-estimating a health statistic, unnecessary stress can suffer patients or policymakers would invest resources on ineffective health programs.

Quantitative studies are the predominant type of research in current medical literature so that reading most medical sciences research papers requires interpreting statistical results. Although students in medical universities are expected to learn how to interpret these results during several courses, it seems that there is still a lack of knowledge in graduates (2). The role of practitioners is vital because they have access to health statistics relating to their field and act as an interpreter and carrier of this information to their patients.

Health statistics are presented in various ways including risk (e.g. frequencies, percentages, and probabilities) or risk reduction (e.g. relative risk reduction, absolute risk reduction, and number needed to treat). These ways have their advantages and disadvantages; for example, it seems that people understand natural frequencies better than percentages (3). In this study, interpreting a diagnostic test result presenting by percentage among members of a social network was evaluated.

## Methods

### Participants

We recruited participants from the followers of personal Instagram accounts on 30 September 2018. Participants were from a diverse range of disciplines but mainly dental and medical students studying in Iran.

### Question

A question used in a previous research article regarding a positive screening result presented in percentage format (4) was used. I translated and simplified it and put it on the “Stories” section of five Instagram accounts using a “Question Sticker”. Stories are temporary posts which disappear after 24 hours and participants cannot see each other’s responses. The question and its answer are described in Box 1 and Box 2, respectively.

#### Box 1. The question asked in “Stories”.

*The first story*

One simple question:

About 1% of babies born in a population have Down’s syndrome. In a screening test, if the baby actually has Down’s syndrome, there is a 90% chance that the result will be positive. If the baby does not have Down’s syndrome, there is a 1% chance that the test will be positive.

*The second story*

A pregnant woman has given this test and the result has been positive. How likely is that her baby will have Down’s syndrome?

Can you solve this question?

#### Box 2. The answer to the question.

If 10,000 pregnant women were tested, we would expect 100 (1% of 10 000) to have babies with Down’s syndrome.

Of these 100 babies with Down’s syndrome, the test result would be positive for 90 (90% of 100) and negative for 10.

Of the 9900 unaffected babies, 99 (1% of 9900) will also have a positive test, and 9801 will have a negative test result.

So, out of the 10,000 pregnant women tested, we would expect to see 189 (90 + 99) positive test results. Only 90 of these actually have babies with Down’s syndrome, which is 47.6%.

Therefore, 47.6% of pregnant women who have a positive result from the test would actually have a baby with Down’s syndrome.

The correct answer to this question is 47.6%. Positive predictive value (PPV) must be calculated to solve it. The original article proposes the solution method described in Box 2. It is worth noting that there is another method using conditional probability (P (Down’s syndrome | +)) (5).

Answers ranging from 45 to 50 were considered as correct.

### Data collection

Data were collected from five personal Instagram accounts with 3532 followers in total. The collected information included the exact calculated number, educational status (undergraduate or graduated) and field of study. Field of the study was divided into four categories: dentistry, medicine, other medical, and non-medical. The non-medical category was used as control. To avoid duplication of the results, account names were also checked.

### Data analysis

The results were analysed using R version 3.4.4 on Microsoft Windows 10. The Pearson χ2 test was used to compare groups by field of study and educational status.

## Results

Overall, 121 people answered the question (52.9% female). The response rate was not calculated as many people may simply skip the stories and without reading the text. The details of the participants are summarised in Table 1.

**Table 1.** Number of participants in each field and their graduation status.

| Field | Undergraduate | Graduated |
| --- | --- | --- |
| Dentistry | 25 | 11 |
| Medicine | 32 | 0 |
| Pharmacy | 3 | 0 |
| Dental prosthesis technician | 1 | 2 |
| Public health | 1 | 2 |
| Occupational therapy | 1 | 0 |
| **Nursing** | 1 | 0 |
| **Health information technology** | 0 | 1 |
| **Midwifery** | 1 | 1 |
| **Laboratory sciences** | 0 | 1 |
| **Electrical engineering** | 10 | 2 |
| **Computer engineering** | 7 | 3 |
| **Architecture** | 1 | 2 |
| **Biotechnology** | 1 | 0 |
| **Chemistry** | 1 | 1 |
| **Civil engineering** | 1 | 1 |
| **Economics** | 0 | 1 |
| **Industrial engineering** | 2 | 0 |
| **Law** | 1 | 0 |
| **Mathematics** | 1 | 0 |
| **Physics** | 0 | 1 |
| **Physical education** | 1 | 0 |
| **Psychology** | 1 | 0 |

Figure 1 shows the correct and incorrect response by field and educational status. The majority of responses (52.1%) were between 0-10% and 90-100%. Overall, 63.6% of the participants did not provide a correct answer. Dental graduates provided the least correct answers ratio among all groups (90.9%). The highest and the lowest correct answer ratios were seen in non-medical and dentistry groups, with 52.6% and 22.2%, respectively (P = 0.09, χ2 = 6.464, df = 3). Only non-medical undergraduates had a correct answer ratio over 50%; however, the difference between undergraduates was not statistically significant (P = 0.05, χ2 = 7.570, df = 3). Undergraduates showed a more favourable performance than graduates with 41.8% correct answers compared to 20.0% (P = 0.03, χ2 = 4.616, df = 1).

**Figure 1.**
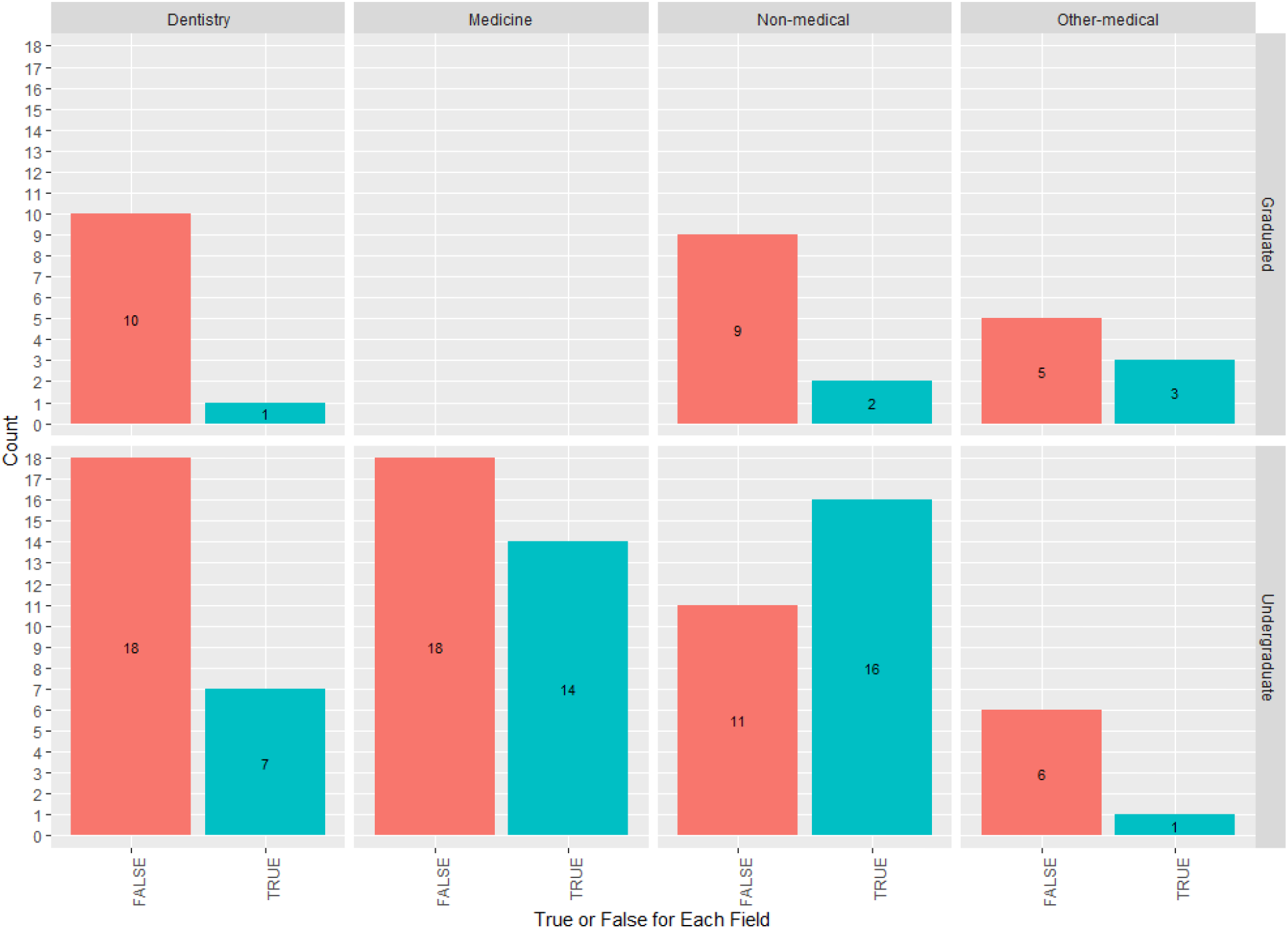
Results of responses to the question, by field of study.

## Discussion

This study indicated that the status of interpretation of statistics among health care professionals is not favourable, especially among participants with dental background. Participants outside the medical field provided the best results. Furthermore, undergraduates provided better results than graduates did. This may demonstrate that graduation has a negative effect on the statistical interpretation ability as graduates deal less with health statistics in their daily practices.

In the study which the question of this study was derived from, midwives and gynaecologists had the lowest percentage of correct answers among other groups (4). However, the results of participants with medical sciences backgrounds and overall results were lower than our study. Still, it seems necessary to improve this situation among students of medical universities in Iran.

Most responses were either very high or very low. Two incorrect answers of 1.0% and 99.0% were the most frequent answers. Respondents who answered 1.0% just used the base rate of Down’s syndrome in the population and those who reached 99.0% considered the conditional probability of observing a positive test in a child without Down’s syndrome. This may show that these participants have not completely understood the concept of PPV and how to calculate it.

### Limitations

Online tests can simplify the study design and save time and other resources. However, controlling the study conditions such as time and use of other sources to answer the question may be a concern. Yet the results of this study can be used to raise awareness about the problem and to design studies of higher quality and at a larger scale.

## Conclusion

This study showed that the statistical interpretation of the medical professionals is lower than the non-medical professionals and graduates showed less favourable results compared to undergraduates. Based on these results, more attention should be paid to the interpretation of medical statistics in medical universities of Iran and the educations should be continued after graduation.

## Data Availability

The data are available as the link below.

https://doi.org/10.6084/m9.figshare.7152659

## Acknowledgments

The author would like to thank Pouria Iranparvar, Hemn Rasouli, Soroush Sadr, and Melika Mohammadi for their help in data collection.

## Notes

### Competing Interest Statement

The authors have declared no competing interest.

### Funding Statement

No external funding was received.

### Author Declarations

Shahid Beheshti Dental School

